# Hepatitis E virus immunoglobulin G seropositivity and kidney dysfunction among United States adults

**DOI:** 10.64898/2026.09.15.26363188

**Authors:** Alara Abu Saadeh, Houssam Eddine Youcefi, Mohamad Ali Kalaji, Layla Alkhuder, Yaman Alsukari, Mahmoud Alothman Agha, Anas Sermani

**Affiliations:** Bahçeşehir University School of Medicine, Istanbul, Turkey; Division of Hematology and Oncology, Department of Internal Medicine, American University of Beirut, Beirut, Lebanon; American University of Beirut, Beirut, Lebanon; Department of Internal Medicine, Damascus Hospital, Damascus, Syria; RAK Medical and Health Sciences University, Ras Al Khaimah, United Arab Emirates

## Abstract

**Background:** Hepatitis E virus infection is associated with renal manifestations in selected clinical populations. Whether serologic evidence of previous exposure is associated with kidney dysfunction in the general population remains uncertain.

**Methods:** We conducted a cross-sectional analysis of six National Health and Nutrition Examination Survey cycles from 2009–2018 and August 2021–August 2023. Adults aged ≥20 years with valid hepatitis E virus immunoglobulin G levels, serum creatinine levels, urinary albumin-to-creatinine ratios, and covariate data were included. The outcomes included a reduced estimated glomerular filtration rate (<60 mL/min/1.73 m²), albuminuria (albumin-to-creatinine ratio ≥30 mg/g [3.39 mg/mmol]), and their composite. Survey-weighted logistic regression models were unadjusted, adjusted for age and sex, and additionally adjusted for demographic and clinical covariates.

**Results:** Among 30,428 adults, 2,911 were seropositive; the weighted seroprevalence was 8.29%. The seropositive participants were older than the seronegative participants were (weighted mean, 60.7 versus 46.5 years). Seropositivity was associated with all three binary outcomes in unadjusted analyses, but the estimates approached the null after adjustment for age and sex. The fully adjusted odds ratios were 0.99 (95% confidence interval, 0.85–1.16) for a reduced filtration rate, 1.05 (0.91– 1.22) for albuminuria, and 1.05 (0.93–1.19) for composite kidney dysfunction. In a secondary analysis, seropositivity was associated with a 1.57 mL/min/1.73 m² higher adjusted mean filtration rate (95% confidence interval, 0.71–2.43).

**Conclusions:** Hepatitis E virus immunoglobulin G seropositivity was not significantly associated with binary kidney outcomes after adjustment. Attenuation occurred principally after adjustment for age and sex. These findings do not establish the absence of an association and do not address kidney injury during active or persistent infection.

## Introduction

Hepatitis E virus (HEV) is a cause of acute viral hepatitis worldwide and has been associated with extrahepatic manifestations, including renal disease [1]. Reported renal manifestations include membranoproliferative glomerulonephritis, IgA nephropathy, membranous nephropathy, and abnormalities associated with cryoglobulinemia [2]. Severe glomerular disease has been described in immunocompetent individuals, and renal involvement has been reported among solid-organ transplant recipients with acute or chronic HEV infection [3,4].

The mechanisms underlying HEV-associated renal injury remain incompletely understood. The identification of HEV protein–antibody complexes in glomerular deposits supports a possible immune complex-mediated mechanism [5]. However, anti-HEV immunoglobulin G (IgG) indicates previous exposure and does not distinguish resolved infection from ongoing viral replication. Assessment of active or persistent infection requires virological testing rather than IgG serostatus alone [6].

In the United States, HEV IgG seropositivity increases with age [7], while reduced kidney function is also more common among older individuals [8]. Studies examining HEV and renal disease have included case reports and selected clinical populations, including patients with glomerular disease and transplant recipients [2–4,9]. Population-based analyses of other hepatotropic viruses, including hepatitis C virus (HCV), have illustrated the use of nationally representative data to examine associations with kidney dysfunction [10]. Whether HEV IgG seropositivity is associated with kidney dysfunction after accounting for demographic and clinical differences remains uncertain.

We evaluated this association among U.S. adults using six National Health and Nutrition Examination Survey (NHANES) cycles from 2009–2018 and August 2021–August 2023. We examined the reduced estimated glomerular filtration rate (eGFR), albuminuria, and a composite of these outcomes, with continuous eGFR as a secondary outcome. We hypothesized that seropositive adults would have a greater burden of kidney dysfunction, while recognizing that age and other participant characteristics could influence the observed association.

## Materials and methods

### Study design and data source

We conducted a cross-sectional analysis of NHANES, a series of surveys conducted by the National Center for Health Statistics (NCHS), Centers for Disease Control and Prevention. NHANES uses a complex, multistage probability sampling design to represent the civilian, noninstitutionalized U.S. population and combines household interviews, physical examinations, and laboratory measurements [11,12].

We pooled six cycles with available anti-HEV IgG measurements: 2009–2010, 2011–2012, 2013– 2014, 2015–2016, 2017–2018, and August 2021–August 2023. We did not include the combined 2017–March 2020 prepandemic file because participants from the standalone 2017–2018 cycle were already included. The pooled analysis therefore represents the included survey periods, with a gap between 2018 and August 2021.

### Ethics statement

NHANES protocols were approved by the NCHS Research Ethics Review Board, and participants provided written informed consent. This analysis used deidentified, publicly available data and did not require additional institutional review. The publicly available NHANES datasets were accessed for this study between 12 June 2026 and 25 August 2026. The authors had no access to information that could identify individual participants at any stage of data acquisition or analysis.

### Study population

Adults aged ≥20 years were eligible if they had valid HEV IgG results, serum creatinine measurements, and available urinary albumin and creatinine measurements. Participants with missing or indeterminate HEV IgG results were excluded. We also excluded participants who reported dialysis during the previous 12 months (KIQ025) and those missing any covariate required for the fully adjusted model. All the primary regression models use the same complete-case sample. Study size was determined by the number of eligible participants with complete data; no a priori sample size calculation was performed.

### Exposure definition

The exposure was HEV IgG serostatus, recorded as LBDHEG. Participants were classified as seropositive or seronegative according to the released NHANES laboratory coding, with equivocal, indeterminate, or missing results excluded. The NHANES documentation describes an enzyme immunoassay for anti-HEV IgG; the assay is identified as abia HEV IgG in the 2017–2018 and August 2021–August 2023 documentation, with the latter noting its former designation as DS-EIA-ANTI-HEV-G [13,14]. Seropositivity was interpreted as evidence of previous exposure, not as confirmation of active infection.

### Kidney outcomes and serum creatinine harmonization

Kidney outcomes were derived from serum creatinine levels and the urinary albumin-to-creatinine ratio (ACR). To account for changes in creatinine assays, values from 2009–2010 through 2015– 2016 were transformed to the 2017–2018 Roche Cobas 6000 scale using the NCHS bridging equation [15]:

Harmonized creatinine (mg/dL) = 0.9515 × released creatinine (mg/dL) + 0.06608.

Creatinine in mg/dL can be converted to μmol/L by multiplying by 88.4. The equivalent equation in μmol/L is harmonized creatinine = 0.9515 × released creatinine + 5.8415. No additional adjustment was applied to the 2017–2018 or August 2021–August 2023 values [15,16]. We calculated the eGFR from harmonized creatinine using the 2009 Chronic Kidney Disease Epidemiology Collaboration (CKD-EPI) creatinine equation [17,18].

Reduced eGFR was defined as <60 mL/min/1.73 m². Urinary ACR was calculated from urinary albumin and creatinine concentrations, and albuminuria was defined as an ACR ≥30 mg/g (3.39 mg/mmol) [19]. The three binary outcomes were reduced eGFR, albuminuria, and composite kidney dysfunction, defined as either reduced eGFR or albuminuria. Continuous eGFR was a secondary outcome. Because measurements were obtained at a single examination, these outcomes indicate kidney dysfunction rather than confirmed chronic kidney disease.

### Covariates

The fully adjusted model included age, sex, race, body mass index (BMI), diabetes status, hypertension status, cardiovascular disease status, and previous stroke status. Age and BMI were modeled continuously. Race was classified as non-Hispanic Black versus all other racial and ethnic categories using RIDRETH1 for 2009–2010 and RIDRETH3 for subsequent cycles; category 4 identified non-Hispanic Black participants in terms of both variables. We refer to this binary variable as non-Hispanic Black versus other races and ethnicities below.

Diabetes and hypertension were based on self-reported physician diagnoses. Cardiovascular disease was defined as a self-reported history of congestive heart failure, coronary heart disease, angina, or myocardial infarction. Previous stroke was based on self-reported physician diagnosis.

### Survey weighting

The analyses incorporated sampling weights, masked primary sampling units, and strata. We used the 2-year Mobile Examination Center weight (WTMEC2YR) for 2009–2010 through 2017–2018 and the 2-year phlebotomy weight (WTPH2YR) for August 2021–August 2023. Phlebotomy weight accounts for nonresponse in blood collection and is recommended for analyses involving blood analytes in that cycle [14,16]. Each cycle-specific weight was divided by six to construct the pooled analysis weight [20]. Strata were nested within the survey cycle to distinguish identically numbered strata across cycles. The final survey design had 93 degrees of freedom.

### Statistical analysis

Participant characteristics were summarized by HEV IgG serostatus. Approximately symmetric continuous variables are reported as survey-weighted means with standard errors (SEs), and categorical variables are reported as survey-weighted percentages. Because urinary ACR was right skewed, it was reported as a survey-weighted median and interquartile range (IQR). Between-group comparisons used survey-weighted t tests for means, Rao–Scott adjusted chi-square tests for categorical variables, and a design-based Wilcoxon rank test for ACR. For the three binary kidney outcomes, unweighted event counts and denominators were reported alongside weighted prevalence estimates.

Associations between HEV IgG seropositivity and each binary outcome were estimated using survey-weighted logistic regression and are expressed as odds ratios (ORs) with 95% confidence intervals (CIs). Seronegative participants composed the reference group. Model 1 was unadjusted; Model 2 was adjusted for age and sex; and Model 3 was additionally adjusted for race and ethnicity, BMI, diabetes, hypertension, cardiovascular disease, and previous stroke. For presentation of the full composite-outcome model, age was expressed per 10-year increase. The survey cycle was incorporated into the variance design but was not included as a regression covariate.

Continuous eGFR was analyzed using survey-weighted linear regression with Model 3 covariates. In a prespecified subgroup analysis, the survey design was restricted to HEV IgG-seropositive participants, and composite kidney dysfunction was regressed on age, sex, race and ethnicity, BMI, diabetes, hypertension, cardiovascular disease, and previous stroke. This analysis assessed covariate associations within seropositive participants; it did not estimate the association of serostatus with kidney dysfunction or test interactions by serostatus. No separate sensitivity analysis was performed.

Analyses were conducted in R version 4.6.1 using the survey package [21]. Tests were two-sided, with P < 0.05 considered to indicate statistical significance. The primary binary outcomes, secondary continuous outcomes, and subgroup results were interpreted together, with attention to effect sizes and confidence intervals.

## Results

### Study population

The pooled dataset contained 61,626 participants. The exclusion of 30,844 participants aged <20 years or with missing or indeterminate HEV IgG results or missing kidney measurements left 30,782 eligible adults. We then excluded 62 participants who reported dialysis during the previous 12 months and 292 with missing covariates, yielding 30,428 participants. Of these, 2,911 (9.57%, unweighted) were HEV IgG seropositive, and 27,517 (90.43%, unweighted) were seronegative. The weighted seroprevalence was 8.29% (SE, 0.32 percentage points). Participant selection is shown in Fig 1. All the primary models used this same complete-case sample.

**Fig 1.**
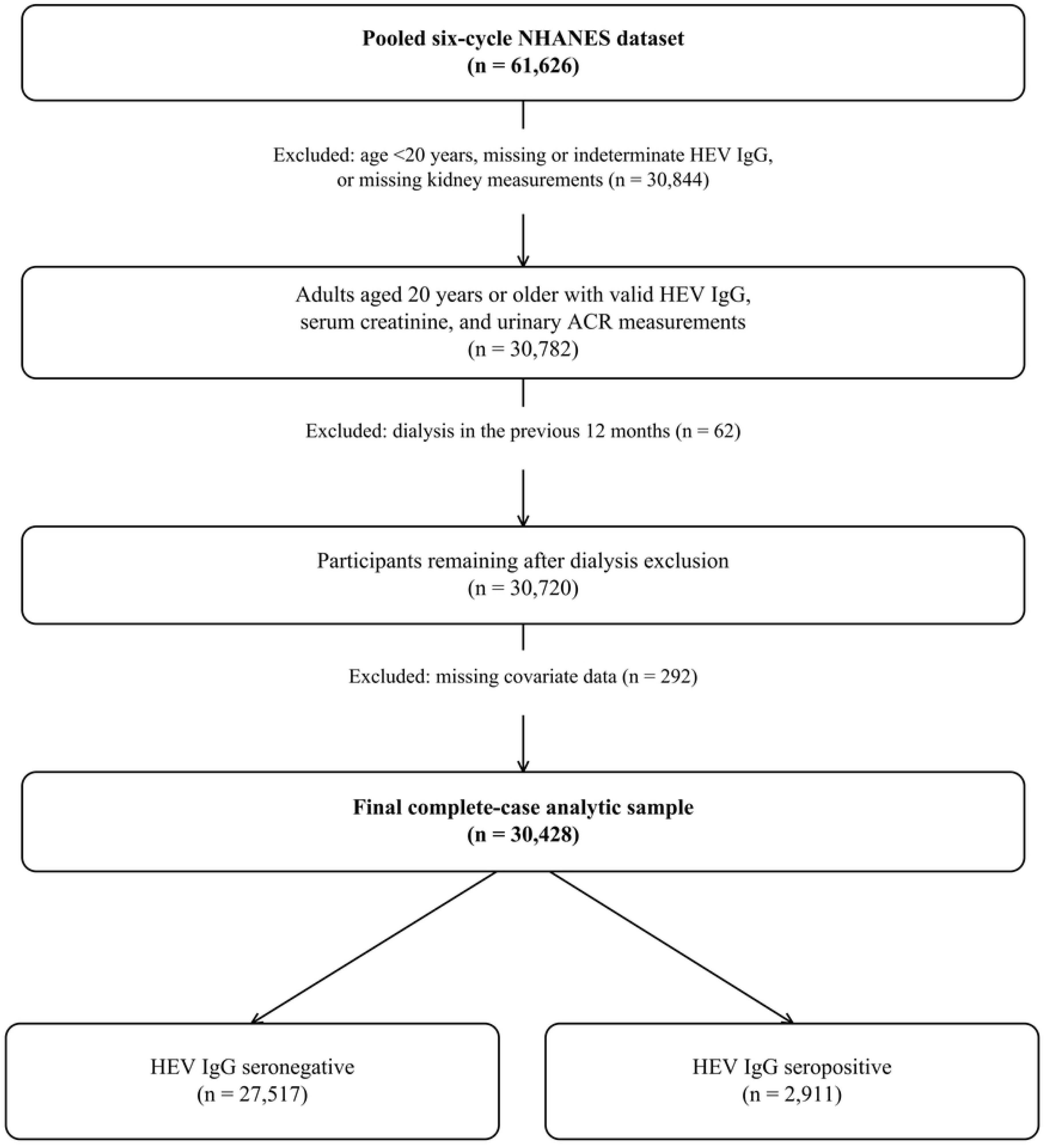
Participant selection flowchart. Sequential exclusions from six National Health and Nutrition Examination Survey cycles (2009–2018 and August 2021–August 2023) to the complete-case sample of 30,428 adults. Counts are unweighted. ACR, albumin-to-creatinine ratio; HEV, hepatitis E virus; IgG, immunoglobulin G.

### Participant characteristics

Table 1 presents participant characteristics by HEV IgG serostatus. Compared with seronegative participants, seropositive participants were older (weighted mean age, 60.7 versus 46.5 years) and had higher weighted prevalences of diabetes, hypertension, cardiovascular disease, and previous stroke (all P < 0.001). The sex distribution and mean BMI did not differ significantly between the groups.

**Table 1.** Characteristics of participants by hepatitis E virus IgG serostatus.

| Characteristic | Seronegative<br>(n = 27,517) | Seropositive<br>(n = 2,911) | P value |
| --- | --- | --- | --- |
| Age, years, mean (SE) | 46.5 (0.24) | 60.7 (0.45) | <0.001 |
| Male sex, % | 48.3 | 48.2 | 0.895 |
| Non-Hispanic Black race, % | 11.1 | 6.1 | <0.001 |
| BMI, kg/m<sup>2</sup>, mean (SE) | 29.3 (0.09) | 29.3 (0.21) | 0.877 |
| Diabetes, % | 9.6 | 15.0 | <0.001 |
| Hypertension, % | 30.5 | 43.8 | <0.001 |
| Cardiovascular disease, % | 6.2 | 12.2 | <0.001 |
| Previous stroke, % | 2.6 | 4.5 | <0.001 |
| eGFR, mL/min/1.73 m<sup>2</sup>, mean (SE) | 93.8 (0.30) | 82.4 (0.63) | <0.001 |
| Urinary ACR, mg/g, median (IQR) | 6.6 (4.4–11.8) | 8.0 (5.3–15.5) | <0.001 |
| Equivalent in mg/mmol | 0.75 (0.50–1.33) | 0.90 (0.60–1.75) |  |
| Reduced eGFR, weighted % (n/N) | 6.3 (2,189/27,517) | 15.0 (491/2,911) | <0.001 |
| Albuminuria, weighted % (n/N) | 9.1 (3,160/27,517) | 13.2 (450/2,911) | <0.001 |
| Composite kidney dysfunction, weighted % (n/N) | 13.6 (4,623/27,517) | 24.1 (796/2,911) | <0.001 |

Values are survey-weighted means (SE), percentages, or medians (IQR). Sample sizes and event counts (n/N) are unweighted. The eGFR was <60 mL/min/1.73 m<sup>2</sup>, the serum concentration of albuminuria was ≥30 mg/g (3.39 mg/mmol), and composite kidney dysfunction was an outcome. ACR values in mg/mmol are unit conversions of the reported values in mg/g, rounded to two decimals. P values are from survey-weighted t tests for means, Rao–Scott adjusted chi-square tests for categorical variables, and a design-based Wilcoxon rank test for ACR. ACR, albumin-to-creatinine ratio; BMI, body mass index; eGFR, estimated glomerular filtration rate; HEV, hepatitis E virus; IgG, immunoglobulin G; IQR, interquartile range; SE, standard error.

Seropositive participants had lower unadjusted mean eGFRs (82.4 versus 93.8 mL/min/1.73 m²) and higher median ACRs (8.0 versus 6.6 mg/g; 0.90 versus 0.75 mg/mmol). The weighted prevalence rates of reduced eGFR, albuminuria, and composite kidney dysfunction were 15.0%, 13.2%, and 24.1%, respectively, among seropositive participants and 6.3%, 9.1%, and 13.6%, respectively, among seronegative participants (all P < 0.001). The corresponding unweighted event counts and denominators are provided in Table 1.

### Associations between seropositivity and kidney outcomes

Table 2 presents the associations between HEV IgG seropositivity and each binary kidney outcome across the three models. Unadjusted ORs were 2.65 (95% CI, 2.31–3.03) for reduced eGFR, 1.53 (1.31–1.78) for albuminuria, and 2.02 (1.79–2.27) for composite kidney dysfunction (all P < 0.001).

**Table 2.** Associations between hepatitis E virus IgG seropositivity and kidney outcomes.

| **Outcome** | **Model** | **OR (95% CI)** | **P value** |
| --- | --- | --- | --- |
| Reduced eGFR | 1 | 2.65 (2.31–3.03) | <0.001 |
| Reduced eGFR | 2 | 0.97 (0.84–1.12) | 0.685 |
| Reduced eGFR | 3 | 0.99 (0.85–1.16) | 0.886 |
| Albuminuria | 1 | 1.53 (1.31–1.78) | <0.001 |
| Albuminuria | 2 | 1.00 (0.86–1.16) | 0.970 |
| Albuminuria | 3 | 1.05 (0.91–1.22) | 0.496 |
| Composite kidney dysfunction | 1 | 2.02 (1.79–2.27) | <0.001 |
| Composite kidney dysfunction | 2 | 1.01 (0.90–1.14) | 0.834 |
| Composite kidney dysfunction | 3 | 1.05 (0.93–1.19) | 0.404 |

ORs compare seropositive with seronegative participants (reference). Model 1 was unadjusted. Model 2 was adjusted for age and sex. Model 3 was additionally adjusted for non-Hispanic black individuals versus other races and ethnicities and for BMI, diabetes status, hypertension status, cardiovascular disease status, and previous stroke status. All the models used survey-weighted logistic regression for the same 30,428 participants. The eGFR was <60 mL/min/1.73 m<sup>2</sup>, the serum concentration of albuminuria was ≥30 mg/g (3.39 mg/mmol), and composite kidney dysfunction was an outcome. ACR, albumin-to-creatinine ratio; BMI, body mass index; CI, confidence interval; eGFR, estimated glomerular filtration rate; IgG, immunoglobulin G; OR, odds ratio.

After adjustment for age and sex, the estimates approached null: reduced eGFR, OR 0.97 (95% CI, 0.84–1.12; P = 0.685); albuminuria, OR 1.00 (0.86–1.16; P = 0.970); and composite kidney dysfunction, OR 1.01 (0.90–1.14; P = 0.834). Further adjustment produced little change. The fully adjusted ORs were 0.99 (0.85–1.16; P = 0.886), 1.05 (0.91–1.22; P = 0.496), and 1.05 (0.93–1.19; P = 0.404), respectively. These fully adjusted estimates are displayed in Fig. 2.

**Fig 2.**
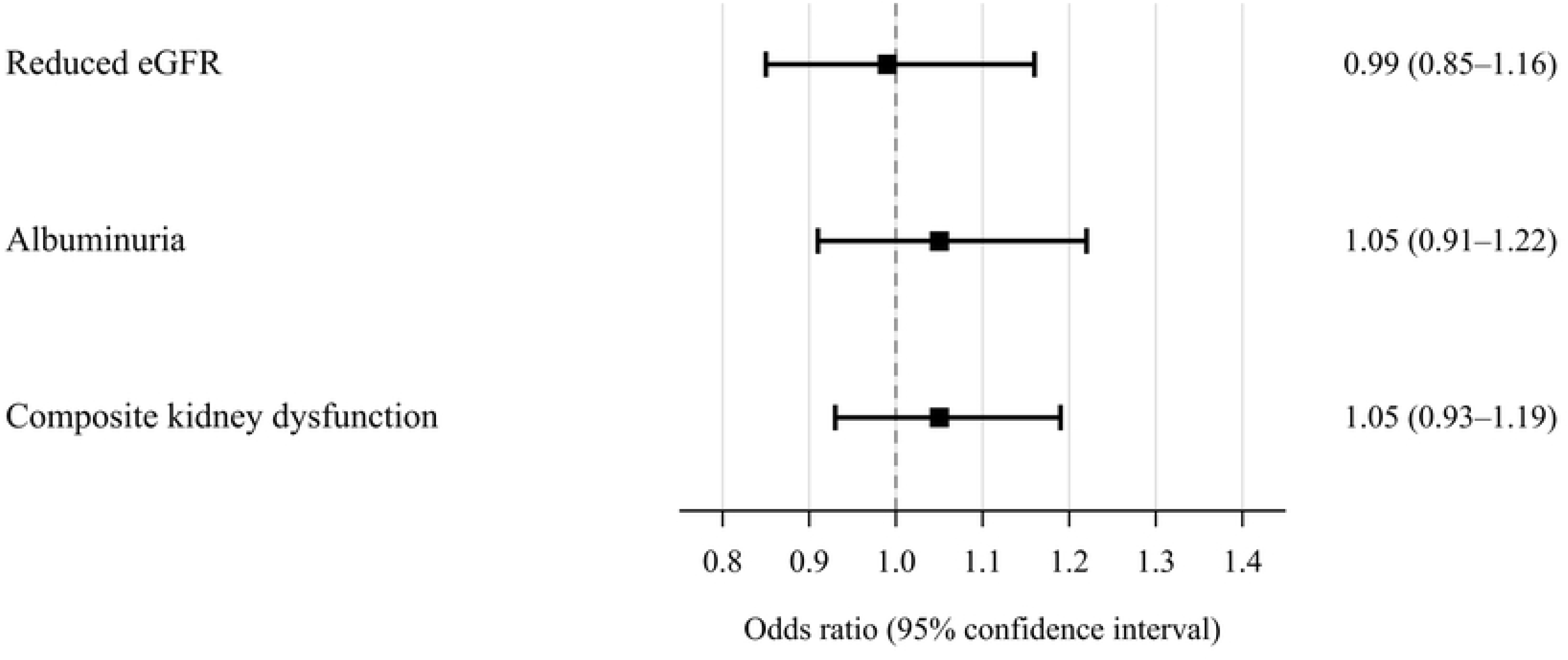
Fully adjusted associations between hepatitis E virus IgG seropositivity and kidney outcomes. Odds ratios and 95% confidence intervals are from survey-weighted logistic regression adjusted for age, sex, non-Hispanic Black status versus other races and ethnicities, body mass index, diabetes status, hypertension status, cardiovascular disease status, and previous stroke status. Seronegative participants composed the reference group; n = 30,428 for each model. The eGFR was <60 mL/min/1.73 m², the serum concentration of albuminuria was ≥30 mg/g (3.39 mg/mmol), and composite kidney dysfunction was an outcome. ACR, albumin-to-creatinine ratio; eGFR, estimated glomerular filtration rate; IgG, immunoglobulin G.

Table 3 presents all covariate ORs from the fully adjusted composite-outcome model. Older age, higher BMI, non-Hispanic Black race, diabetes, hypertension, cardiovascular disease, and previous stroke were associated with higher odds of composite kidney dysfunction. Male sex was associated with lower odds than female sex was. These are mutually adjusted associations within the specified model.

**Table 3.**
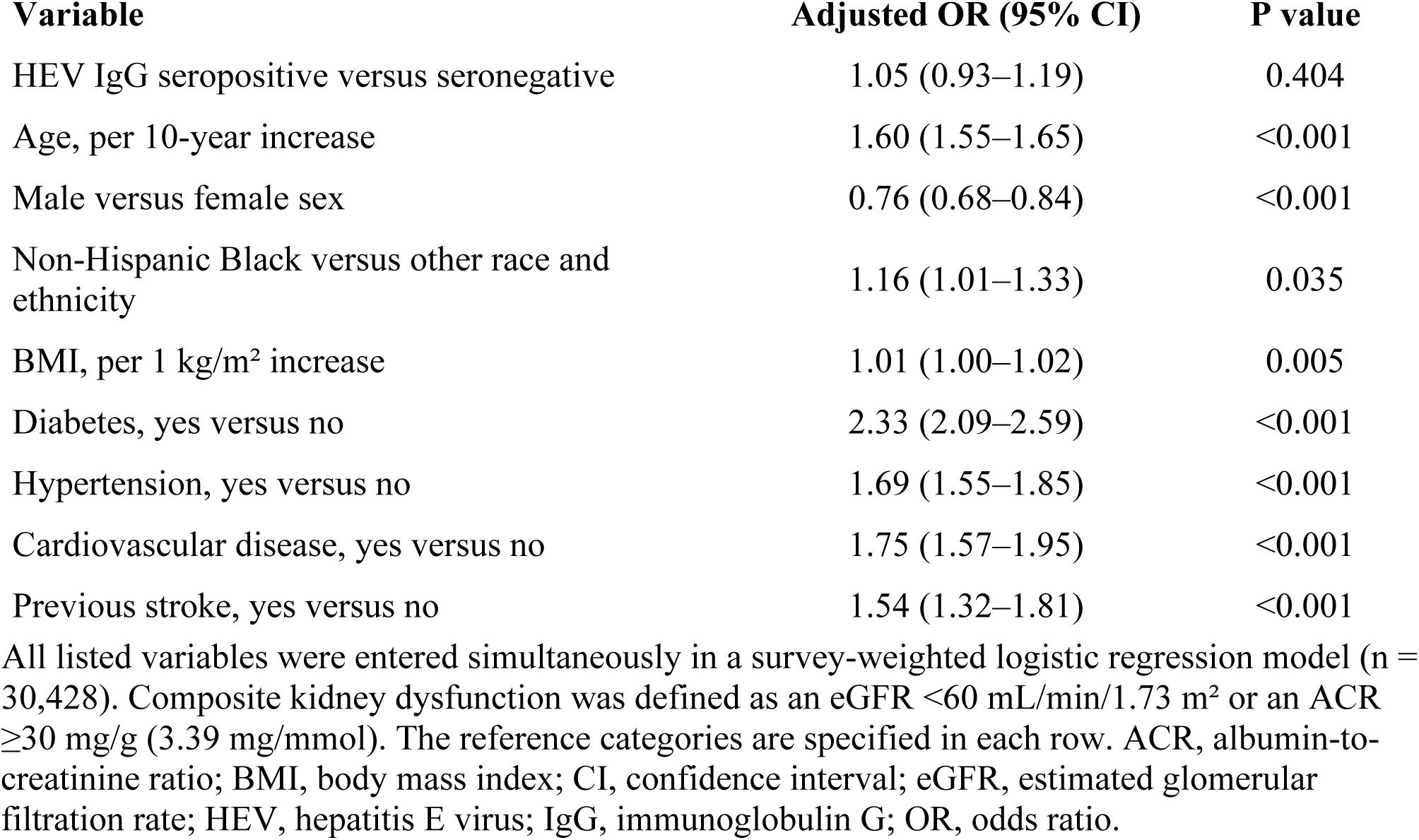
Fully adjusted model for predicting kidney dysfunction among United States adults.

### Secondary analysis of continuous eGFR

In the same complete-case sample, HEV IgG seropositivity was associated with a 1.57 mL/min/1.73 m² higher adjusted mean eGFR (SE, 0.43; 95% CI, 0.71–2.43; P < 0.001). This association was opposite that of the unadjusted comparison in Table 1. The coefficients for the full linear model are provided in Table S1.

### Subgroup analysis among seropositive participants

Among the 2,911 seropositive participants, composite kidney dysfunction was associated with age (OR per 10-year increase, 2.01; 95% CI, 1.74–2.33), diabetes (OR 2.21; 1.59–3.09), hypertension (OR 1.93; 1.43–2.60), cardiovascular disease (OR 1.70; 1.28–2.24), and previous stroke (OR 1.49; 1.04–2.12). The P values were <0.001 for age, diabetes, hypertension, and cardiovascular disease and 0.029 for stroke. Sex, race, ethnicity, and BMI were not significantly associated with the outcome in this subgroup (Table S2).

## Discussion

In this analysis of 30,428 U.S. adults, HEV IgG seropositivity was associated with higher unadjusted prevalence rates and odds of reduced eGFR, albuminuria, and composite kidney dysfunction. All three ORs approached 1 after adjustment for age and sex, with little additional change after adjustment for the remaining covariates. The fully adjusted confidence intervals included 1 for each binary outcome. These results provide no clear evidence of an adjusted association within the models examined, but they do not demonstrate equivalence or exclude smaller associations.

### Interpretation of the adjusted associations

The seropositive participants were approximately 14 years older on average than the seronegative participants were. Previous U.S. studies have likewise reported higher HEV IgG seroprevalence at older ages [7,22], and age is associated with reduced kidney function [8]. The attenuation after adjustment for age and sex is therefore consistent with demographic confounding. Because age and sex were entered together, their separate contributions cannot be determined from these models. Moreover, changes in logistic regression ORs do not quantify the proportion of an association explained by particular covariates. Additional adjustment for clinical comorbidities changed the HEV estimates only slightly.

The secondary association with higher adjusted mean eGFR should be distinguished from the binary-outcome findings. The estimated difference was small, and a shift in the mean eGFR did not correspond to a difference in the odds of an eGFR below 60 mL/min/1.73 m². Its reversal from the unadjusted comparison also makes the result sensitive to the interpretation of covariate adjustment. It does not establish a beneficial effect of previous HEV exposure. The subgroup analysis describes associations within seropositive participants; differences in statistical significance between that subgroup and the full sample cannot establish effect modification.

### Comparison with previous studies

Studies of HEV exposure and glomerular disease have reported differing results. Pischke et al. reported no significant difference in anti-HEV IgG seroprevalence between 108 patients with glomerulonephritis and 108 age- and sex-matched controls [23]. In contrast, El-Mokhtar et al. reported higher seroprevalence among 43 Egyptian patients with glomerulonephritis than among 36 healthy controls [24]. These studies involved selected clinical populations and assessed glomerulonephritis rather than the broad kidney function measures used here. Differences in population, setting, exposure ascertainment, and outcome definition limit direct comparisons. HCV provides a related but distinct comparison. In an NHANES analysis, Chen et al. reported adjusted associations of resolved and chronic HCV infection with kidney disease [10]. Our findings concern HEV IgG serostatus and should not be generalized to other hepatotropic viruses.

Differences in viral persistence, host factors, and the classification of infection may contribute to the differing associations; this study did not directly compare the effects of HEV and HCV. IgG seropositivity does not indicate active HEV infection [6]. Persistent infection has been reported in immunosuppressed transplant recipients [25], and renal abnormalities have been described in transplant recipients with HEV infection [4]. Marion et al. reported worse kidney function among chronically infected transplant recipients with cryoglobulinemia and a reduction in the prevalence of cryoglobulin after viral clearance [26]. Leblond et al. identified HEV ORF2 protein–antibody complexes in glomerular deposits, supporting an immune complex-mediated mechanism in HEV-associated glomerulonephritis [5]. A reported case in an immunocompetent individual revealed that such renal manifestations are not confined to transplant recipients [3]. These clinical observations concern settings that an IgG-based population analysis cannot adequately distinguish.

### Strengths and limitations

Strengths include the large NHANES sample, incorporation of the complex survey design, assessment of complementary kidney outcomes, and use of the same complete-case sample across the primary models. Sequential adjustment revealed that attenuation occurred primarily after the inclusion of age and sex. Creatinine harmonization addresses an identified assay difference, and the confidence intervals provide information about the range of associations compatible with the fitted models.

The cross-sectional design prevents the determination of whether HEV exposure preceded kidney dysfunction and precluded causal inference. IgG serostatus cannot be used to determine infection timing or to distinguish resolved exposure from active or persistent infection [6]. HEV RNA, viral load, and genotype were not evaluated, so the findings do not resolve questions about the renal consequences of active HEV infection.

Kidney measurements were obtained at a single examination. Persistence for at least 3 months is needed to establish chronic kidney disease [19]; therefore, transient abnormalities cannot be distinguished from chronic disease. Differences in the results of residual assays may remain despite the use of creatinine harmonization. The 2009 CKD-EPI equation includes a race coefficient, which may affect outcome classification and limit comparability with newer equations that do not include race. Associations with age, sex, and race also require caution because these characteristics are included in the eGFR equation itself.

The pooled estimates describe the included periods rather than an uninterrupted interval or the current U.S. population. The survey cycle was not included as a regression covariate, and changes in seroprevalence, population composition, or kidney health across periods could leave residual confounding. Age and BMI were modeled as linear terms; nonlinear relationships could also affect adjustment. The binary race and ethnicity variable combines heterogeneous populations and may not adequately capture differences relevant to HEV exposure and kidney health.

Self-reported diagnoses may miss undiagnosed disease and are subject to recall and classification errors. Complete-case selection and the requirement for blood and urine measurements may introduce selection bias; the relatively small number of patients excluded for missing covariates does not establish the absence of bias from other exclusions. Residual confounding by unmeasured factors, including socioeconomic characteristics and other infections, remains possible. Multiple outcomes and subgroup comparisons warrant cautious interpretation, particularly for secondary findings. Finally, the results may not be generalizable to institutionalized adults, dialysis patients, or selected populations with active or persistent HEV infection.

## Conclusions

Among U.S. adults in the included NHANES cycles, HEV IgG seropositivity was not significantly associated with reduced eGFR, albuminuria, or composite kidney dysfunction after adjustment. Attenuation of the crude associations occurred principally after adjustment for age and sex. A small positive association with continuous eGFR does not establish a beneficial effect. Longitudinal studies with virological testing and repeated kidney measurements are needed to examine the renal consequences of active or persistent HEV infection.

## Data Availability

No data were generated by this study. The study used publicly available data from the National Health and Nutrition Examination Survey (NHANES), provided by the National Center for Health Statistics, Centers for Disease Control and Prevention. NHANES datasets and associated documentation are publicly available from the CDC/NCHS NHANES website (https://www.cdc.gov/nchs/nhanes/).

## Acknowledgments

None.

## Supporting information captions

**S1 Table.** Fully adjusted survey-weighted linear regression analysis of continuous eGFR. The analysis included 30,428 adults. Coefficients express adjusted differences in eGFR in mL/min/1.73 m². eGFR, estimated glomerular filtration rate.

**S2 Table.** Factors associated with kidney dysfunction among hepatitis E virus IgG-seropositive participants. The survey-weighted logistic regression analysis included 2,911 adults. Composite kidney dysfunction was defined as an eGFR <60 mL/min/1.73 m² or a urinary ACR ≥30 mg/g (3.39 mg/mmol). ACR, albumin-to-creatinine ratio; eGFR, estimated glomerular filtration rate; IgG, immunoglobulin G.

## Notes

### Competing Interest Statement

The authors have declared no competing interest.

### Author Declarations

The NHANES protocols were approved by the National Center for Health Statistics (NCHS) Research Ethics Review Board, and written informed consent was obtained from all participants. The present study used de-identified, publicly available NHANES data and therefore did not require additional institutional review board approval.

